# A methodological framework for deriving the German food-based dietary guidelines 2024: a multi-objective function including human health, the environment, and observed dietary intakes

**DOI:** 10.1101/2025.11.16.25340354

**Authors:** Anne Carolin Schäfer, Heiner Boeing, Rozenn Gazan, Johanna Conrad, Brecht Devleesschauwer, Kurt Gedrich, Hans Hauner, Anja Kroke, Jakob Linseisen, Micha Limbeck, Stefan Lorkowski, Lukas Schwingshackl, Florent Vieux, Kiran Virmani, Ute Nöthlings, Bernhard Watzl

## Abstract

**Background:** Food-based dietary guidelines (FBDGs) provide recommendations on diets that aim to decrease disease risk and environmental impact, while remaining culturally acceptable for the population. Using mathematical optimization to define such diets, these objectives can be operationalized as components of the objective function.

**Objectives:** To develop a framework for (i) deriving an indicator that quantifies diet-related disease burden and (ii) evaluating different weighting schemes within a three-dimensional diet optimization model.

**Methods:** To address objective (i), disability-adjusted life years (DALYs) from the Global Burden of Disease Study (GBD) and a diet-specific burden estimate based on observational data were transformed into a model-compatible indicator using linear interpolation. To address objective (ii), a linear diet optimization model was developed. It included constraints on nutrients and acceptability, and a three-objective function that minimized disease burden (DALYs), environmental impact (greenhouse gas emissions and land use), and, as a proxy for cultural acceptability, deviation from the observed diet. Forty-two model variations with different weighting schemes were computed and compared regarding component outcomes.

**Results:** In proof-of-concept analyses, the derived health indicator aligned closely with values reported in the literature: 90% (based on GBD) and 99% (based on the observational data) of diet-related DALYs were captured. Among the 42 model variations, strong synergies were observed between health and environmental outcomes. Optimizing exclusively for disease burden or environmental impact resulted in substantial deviations from the observed diet, underscoring the importance of considering cultural acceptability. Model stability improved with the inclusion of all three components.

**Conclusion:** The proposed methodology enables the integration of DALYs and provides insights about various weighting schemes to establish a diet optimization model that minimizes disease burden, environmental impact, and deviation from the observed diet, and serves as the basis to derive FBDG for Germany.

## 1 Introduction

The promotion of public health is a core objective of food-based dietary guidelines (FBDGs) [1–3]. From a physiological perspective, both the fulfilment of nutrient requirements and the intake of health-promoting foods contribute to human health. To identify which foods are associated with an increased or decreased risk of developing non-communicable diseases, epidemiological studies analyze diet-health relations [4,5]. Subsequently, systematic reviews of such studies can be used to inform FBDGs about optimal food intake levels [6], an approach recently utilized in the Nordic Nutrition Recommendations [7].

The food system is responsible for approximately a third of global greenhouse gas emissions (GHGEs) [8]; therefore, minimizing the environmental impact of diets is increasingly recognized as an integral part of developing FBDGs [9–11]. In addition, the successful implementation of FBDGs at the population level requires consideration of current dietary patterns to ensure cultural acceptability [2,12]. Consequently, mathematical optimization has gained acceptance as a novel approach for deriving FBDGs. Such optimization models consist of an objective function that identifies the optimal solution within a space defined by constraints [13]. These models allow for the simultaneous consideration of multiple objectives, including health promotion, respecting population-specific dietary habits, and reducing the environmental impact of food consumption [13,14].

However, while nutrient adequacy is often included in diet optimization models as a determinant of human health, diet-health relations are rarely operationalized. Few studies have considered recommended food group quantities from national FBDGs as constraints [14–16]. Yet, a quantifiable health metric should be assigned to the decision variables to enable the dynamic identification of optimal solutions across the various dimensions. The Global Burden of Disease Study (GBD) provides such a metric by establishing the concept of disability-adjusted life years (DALYs). This concept has been used in many studies, and DALYs for dietary risk factors have been obtained from several systematic reviews of epidemiological studies examining diet-health relations [5,17,18].

So far, diet optimization approaches have most often relied on single-objective functions, typically aiming to minimize deviation from population-specific observed dietary patterns [15,19]. Few studies have attempted to formally integrate multiple sustainability dimensions as a complex multi-objective function [20–24]. Given that the simultaneous minimization of disease burden, environmental impact, and dietary changes is required when developing FBDGs, we operationalized these requirements in the present study as three components in the objective function to enable a flexible trade-off solution [20].

In the context of revising the German FBDG based on an optimization model, a transparent description of the arguments for several methodological decisions is warranted. In this article, we describe the extension of our previous optimization model’s objective function [25] by incorporating health and environmental indicators into the objective function. We present an easy-to-use procedure to derive a dynamic metric for diet-health relations, and systematically investigate the weighting of components within the objective function of an optimization model to identify trade-offs and synergistic effects.

## 2 Methods

### 2.1 Basic model features regarding dietary intake and nutrients

The optimization model applied in this study builds on a previously developed framework, in which we examined the use of a hierarchical food coding system, the mathematical structure of the objective function, and alternative sets of nutrient goals [25]. Decision variables were defined using the European Food Safety Authority’s (EFSA’s) FoodEx2 classification system (version MTX 12.1, exposure hierarchy [26]). The list of decision variables and the matching of food groups to FoodEx2 food codes have been described in detail previously [25].

Dietary intakes for German adults (18–65 years) were retrieved from EFSA’s Comprehensive European Food Consumption Database [27,28]. This data is based on the most recent German National Nutrition Survey II (NVS II) (2005-2007), which utilized two non-consecutive 24-hour dietary recalls from 10,419 men and women [29,30]. Results were weighted for age, sex, residential area, and other socioeconomic factors that may cause deviations from the average adult German population [31]. For database compilation, observed dietary intakes were used as weighting factors to calculate nutrient and environmental indicator values for aggregated food groups, as required for the selected optimization level [13,25].

In view of the absence of evidence defining specific targets for the magnitude of deviation from observed dietary intakes [25], this aspect was incorporated as one component of the objective function. We selected a linear function (Simplex) that minimized relative deviations from observed dietary intakes [25], which led to optimized dietary intakes with changes that were proportional to the habitually consumed food group quantities (i.e. larger changes in drinking water, smaller changes in legumes). The following formula was used in the objective function to minimize the deviation from the observed diet:

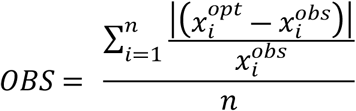

where *n* is the number of food groups (decision variables) from the observed diet, 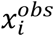 is the observed dietary intake of food group *i,* and 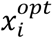 is the optimized quantity of the food group *i.* As described previously [25], acceptability constraints were set at the 5^th^ and 95^th^ percentiles of the observed intakes to avoid unrealistic optimized dietary intakes.

For the nutrient composition and energy content, the German nutrient database Bundeslebensmittelschlüssel (BLS) version 3.02 [32] was used; for free sugars, the LEBTAB database [33] was used. Nutrient goals were based on German dietary reference values (DRVs) for adults (18–65 years, normal weight with moderate physical activity level of 1.4, recommended intakes were used if available) [34] and EFSA’s tolerable upper intake levels for nutrients [35]. Nutrients with limited data quality (iodine, fluoride, copper, and manganese) and vitamin D were excluded from nutrient goals. The applied nutrient goals and mathematical implementation have been described in detail previously [25].

### 2.2 Establishing an indicator for diet-health relations

For modelling diet-health relations, a combination of two different data sources was used. We selected recent data regarding dietary risk factors from the Global Burden of Disease Study 2021 (GBD) [17,36] (S1 Table), and data from a diet-specific burden estimate based on observational data by Schwingshackl et al. [18] (SCH) (S2 Table). Data on the following dietary risk factors were used: a diet low in fruits, vegetables, legumes, whole grains, nuts and seeds, milk (GBD) or dairy (SCH), fish (SCH only), and a diet high in refined grains (SCH only), eggs (SCH only), red meat, processed meat, and sugar-sweetened beverages. The following diet-related disease endpoints were considered: GBD included among others cardiovascular disease, type 2 diabetes mellitus, and cancer, while the SCH analysis included coronary heart disease, stroke, type 2 diabetes mellitus, and colorectal cancer. Food groups that were part of the habitual diet, but for which no DALYs were available (alcoholic beverages, coffee and tea, composite dishes, drinking water, fruit and vegetable juices, potatoes, poultry, seasoning and sauces, spreadable fats, sweets, vegetable fats and oils) were assigned a neutral value (i.e. 0), since the optimization algorithm cannot work with missing values. Although GBD also contains DALYs on nutrient intakes, such as a diet low in calcium or high in salt, we limited our analysis to food-based data.

DALYs originate from complex calculations based on dose-response meta-analyses [4,5,17,18], which result mostly in non-linear risk relations. These non-linear functions were used to derive the theoretical minimum risk exposure level (TMREL), which is defined as the exposure level of a risk factor associated with the lowest disease risk at the population level [37,38]. For example, for the risk factor “diet low in whole grains”, the lowest disease risk was at an intake of 119 g/d of whole grains [18]. Assuming that the observed dietary intakes correspond to diet-related DALYs 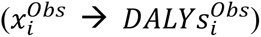, and that optimal dietary intakes (TMREL) correspond to zero DALYs 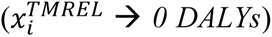, we used linear interpolation to calculate the DALYs corresponding to any food’s *i* dietary intake level being optimized (see formula below and for an example, **Fig 1**, Results section). The linear function used to model DALYs to dietary intakes of each food group was as follows:

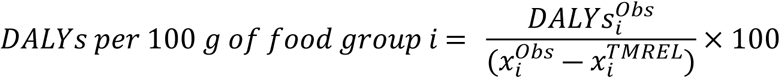

**Fig 1.**
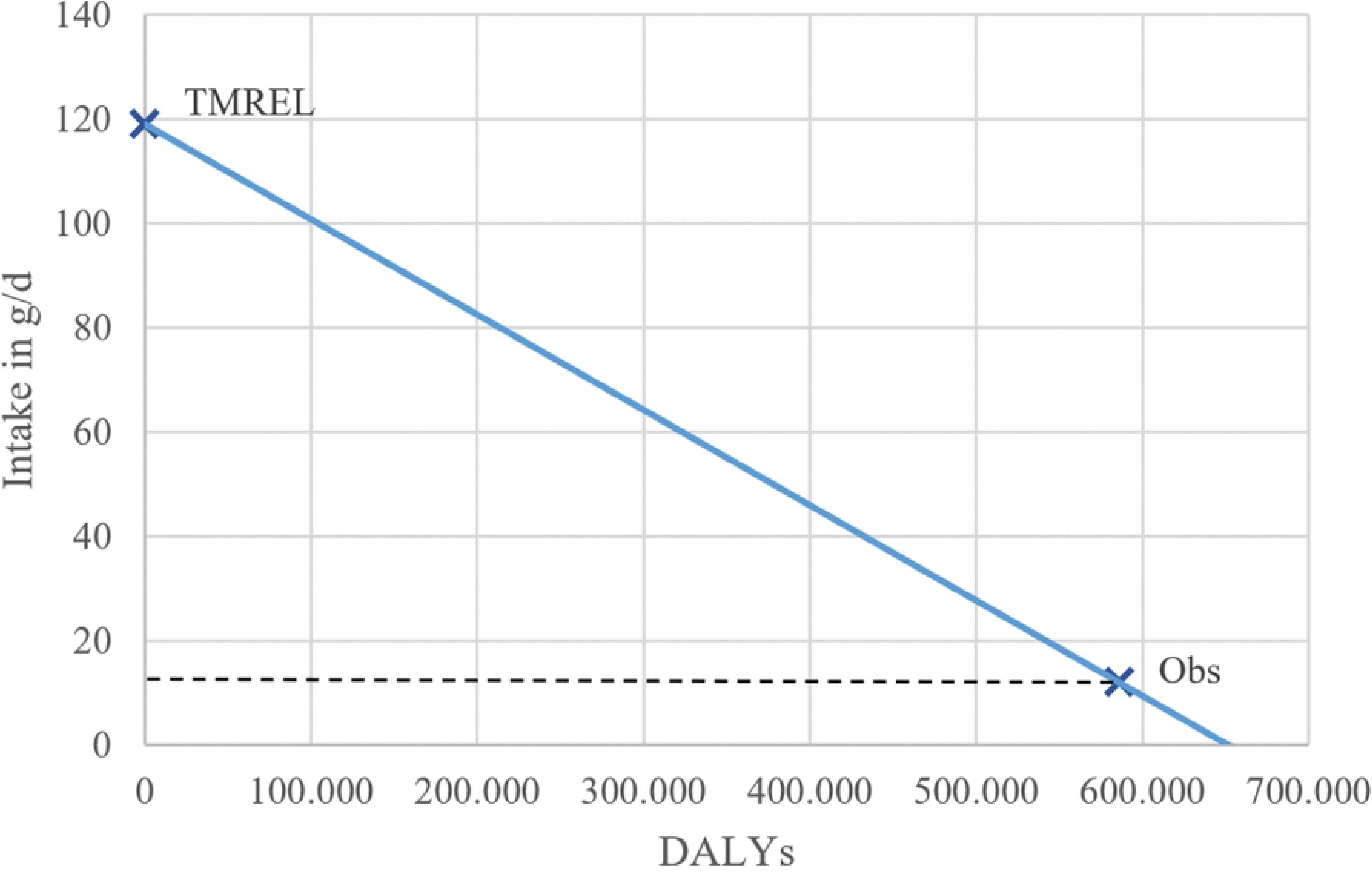
Visualization of the number of DALYs that could be avoided with increasing for whole grain intake (according to SCH data [18]). TMREL = theoretical minimum risk exposure level, Obs = observed, DALYs = disability-adjusted life years.

Data for DALYs were only available for aggregated food groups at higher FoodEx2 levels (e.g. fruits). To obtain data for the food groups on lower aggregation levels (e.g. pome fruits as a decision variable), the DALY value from the respective upper level was assigned to each food group on lower levels using the hierarchical structure of FoodEx2.

As the derivation of DALYs was based on a theoretical ideal (TMREL), the associated optimal consumption values could not be set as specific targets (i.e., in the form of constraints). For example, it is unrealistic, and probably even unnecessary, to set the intake of unfavorable foods to 0 g/d. Instead, it is more realistic to minimize the diet-related disease burden. This procedure leaves room to account for other relevant characteristics of foods, such as nutrient density or environmental impact. The following formula shows the health component (DALYs from GBD and SCH) of the objective function:

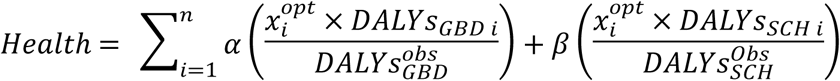

where *α* and *β* are weighting factors (*α* + *β* = 1) for *DALYs*_*GBD* *i*_ and *DALYs*_*Schw* *i*_ (the DALYs according to either GBD or SCH for a food group *i*), and 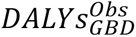 and 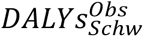 are the total number of DALYs associated with the observed diet from GBD and SCH, respectively. For food groups with positive health impact, no significant further health improvements would be expected beyond TMREL [38]; therefore, the model was adapted so that once this ideal intake level of a food group *i* was achieved, the model could still increase the food group’s quantity, but not reduce further DALYs through that food group.

### 2.3 Environmental indicator data

To integrate environmental impact into the optimization model, a European database of the environmental sustainability of diets, the SHARP Indicators Database (SHARP-ID), was used [39,40]. It comprises a list of 944 FoodEx2 food groups and life cycle assessment data on two indicators, GHGEs in kg CO_2_equivalents/d and land use in m^2^/year/d/kg. The data encompass the whole life cycle of a food from production to transportation and packaging, and, if applicable, food preparations such as energy for cooking, and food waste. To calculate GHGE and land use impacts of food groups aggregated at a higher level (in this study level 3, which provided the decision variables), data were weighted according to dietary intakes of lower-level food groups.

To operationalize reductions in environmental impact, constraints such as planetary boundaries [41], European emission reduction goals [42], progressive reductions (e.g., a 50% reduction of GHGEs) [43,44], or the restriction of food groups related to higher environmental impacts [16] can be used. However, to find the diet with the least environmental impact, the environmental impact should be part of the (multi-) objective function [45,46]. We as well operationalized the environmental impact as one component of our objective function:

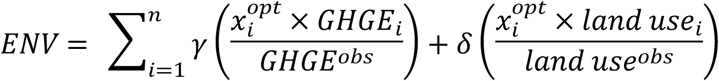

where *γ* and *δ* are weighting factors (*γ* + *δ* = 1) for GHGEs and land use, respectively; *GHGE*_*i*_ and *land use*_*i*_ are the environmental impact for a food group *i*; and *GHGE*^*obs*^ and *land use*^*obs*^ are the total amount of GHGEs and land use in the observed diet, respectively.

### 2.4 Objective function with three components

The objective function aims to minimize *F*_*multi*_; the weighted sum of the three components (diet-health relations [health], environmental impact [ENV], and deviations from observed dietary intakes [OBS]). The mathematical formula of the objective function was as follows:

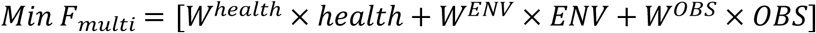

where *W*^*health*^+ *W*^*ENV*^+ *W*^*OBS*^= 1. Technically, the fulfilment of the nutrient goals is also a part of the objective function, but it operates like a constraint as long as the nutrient goals can be reached [25]. It is separately weighted and does not search for a local optimum together with the other three components and therefore is not of interest for the analyses.

To allow the objective function to process each component equally, the components health, ENV, and OBS were standardized (i.e., expressed as a percentage of their observed diet value). Further, to ensure comparability of the health component with the other two components in the objective function, the values of the metric DALYs/100 g food *i* (*DALYs*_*i*_) were transformed into positive values only (transformed indicator *DALYs*_*trans*_(*i*)), while keeping the same relative distances among the negative values (< 0 for *DALYs*_*i*_*, (NEG)*), as well as among the positive values (> 0 for *DALYs*_*i*_*, (POS)*). To achieve this, negative values were transformed to obtain 1 as a new lowest value. Positive values were transformed considering the distance between the lowest positive value and the highest value among negatives values (the value closest to 0) from *DALYs*_*i*_. To transform null values, the positions of the original null value between the highest negative and lowest positive *DALYs*_*i*_value were identified, and transformed values were assigned based on that range.

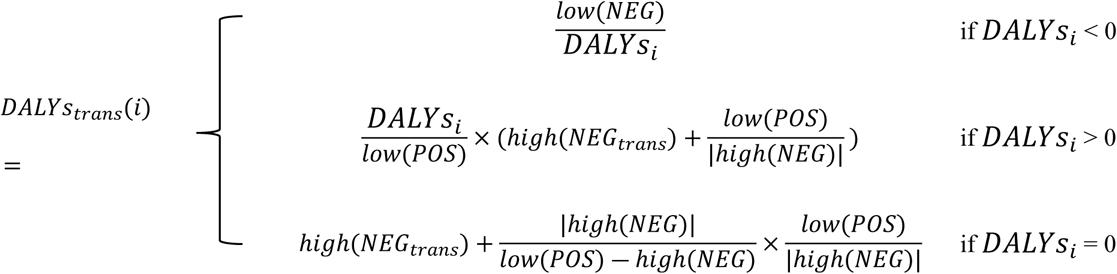

where *low*(*NEG*) is the lowest value from *DALYs*_*i*_, *low*(*POS*) is the lowest positive value, *high*(*NEG*_*trans*_) is the highest transformed, originally negative, DALY values, and *high*(*NEG*) is the highest value (closest to 0) among negatives values from *DALYs*_*i*_.

### 2.5 Analysis

First, as proof-of-concept, derived indicators of diet-health relations (DALYs/100 g of food group *i*) were used to calculate reductions in the disease burden (*DALYs*_*red*_) of a diet consisting of (i) the ideal intake values (TMREL) and (ii) the observed intakes *x* of each food group *i*:

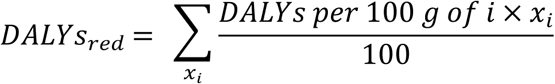

We hypothesized that these health indicators would be suitable for use in the optimization algorithm, as well as to calculate *DALYs*_*red*_ of the optimized diets, if the number of DALYs for both tests was comparable to those in the literature.

Secondly, to study the impact of different weightings of the three components in the objective function, one of the components was fixed to around one-third of the total weighting (35%), while the weightings of the other two components were increased and decreased incrementally by 5%. GHGEs and land use, as well as DALYs from GBD and SCH, were equally weighted within their component of the objective function. Acceptability and nutrient constraints were the same across all models. To study how the models performed using the different weightings for the three components, we compared the following outcomes:

- the relative reduction of DALYs (mean of SCH and GBD data, output of the model which includes data from food groups with neutral values for DALYs, see Chapter 2.2);
- the relative reduction of environmental impact (mean of GHGEs and land use);
- and the average relative change per decision variable from observed to optimized dietary intake quantities (see previously reported formula [26]).

To investigate the influence of the correlations between health, environment and observed diet, as reflected by their indicators, on optimization results, we ran Pearson’s product-moment correlation tests using the R function cor.test. We ran a correlation analysis between the mean of GBD and SCH and the mean of GHGEs and land use to analyze the relation between health and the environment. Next, we ran a correlation analysis between those means and mean dietary intakes to assess how health and environmental factors were associated with the observed diet. Finally, we calculated the correlation between GBD and SCH indicators and GHGEs and land use.

Lastly, sensitivity analyses were conducted to test the robustness of the results regarding the weighting of the components and their indicators. In previous analyses, each indicator (GHGE and land use; GBD and SCH) had a weighting of 50% and the health and environmental results were averaged across indicators (Chapter 3.2). First, we calculated one-dimensional models, with each minimizing only one component (*W*^*Health*^, *W*^*ENV*^, or *W*^*OBS*^= 1). Second, we studied the impact of different weightings of the indicators of the components: GBD and SCH for health, and GHGEs and land use for the environment. In both a one-dimensional model (*W*^*Health*^ or *W*^*ENV*^ = 1) and a multi-dimensional model (*W*^*Health*^ = 0.35, *W*^*ENV*^ = 0.35, and *W*^*OBS*^= 0.3), we tested variations in which both health- and environment-related indicators were included equally, and variations in which only one indicator was selected at a time. Although other combinations of weighting schemes are possible, we decided to test the most diverse.

The models were run in R version 4.1.3 [47], with a custom package using the ROI (R Optimization Infrastructure) package version 0.3-3 [48] and lpsolve [49]. The data were stored and managed with MySQL Workbench version 8.0.

## 3 Results

### 3.1 Operationalizing diet-health relations: Testing the derived indicators

In our model, DALYs constituted the health component in the objective function and were composed of two indicators from two independent sources, as described in Table 1. The first two columns show the originally published DALYs (*DALYs*^*Obs*^) from GBD [36] and SCH [18]. These sources largely agree on the magnitude of DALYs associated with too high or too low consumption of particular food groups, except for nuts and seeds, sugar-sweetened beverages, and red meat.

**Table 1:**
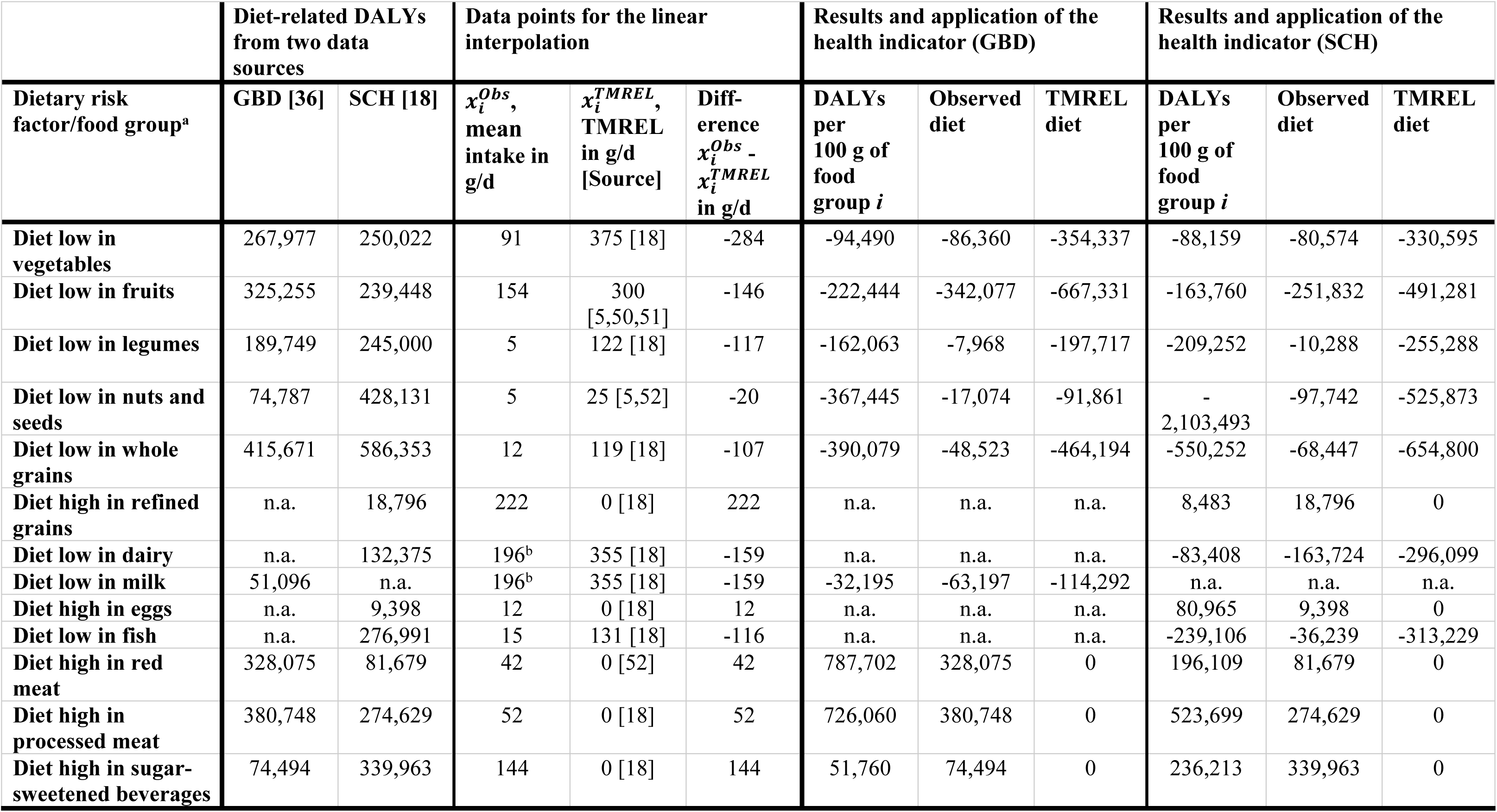
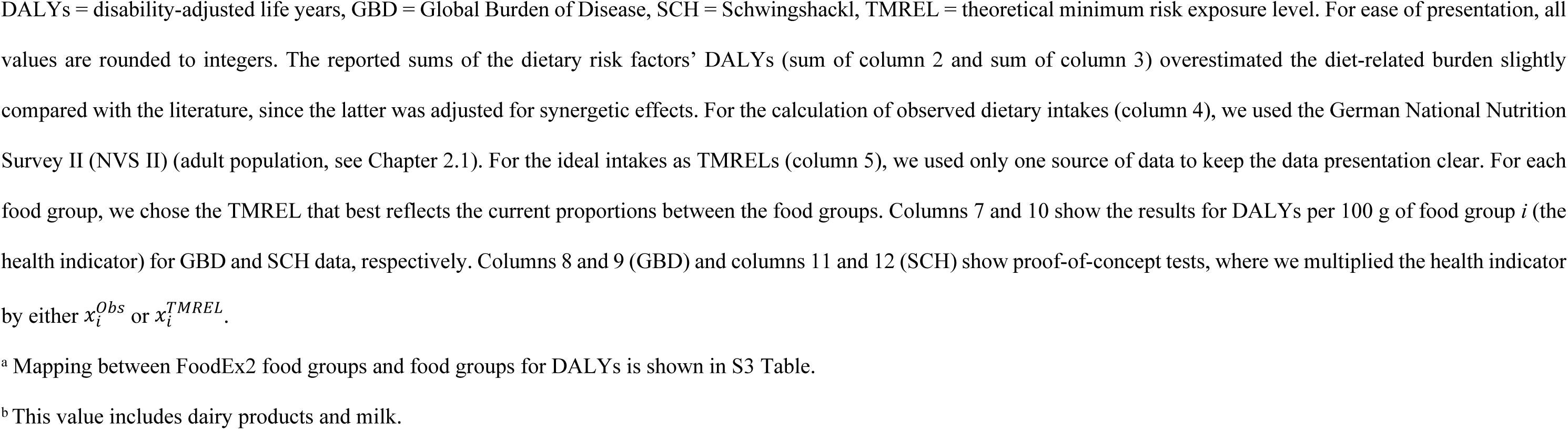
Total DALYs associated with too high or too low consumption of different food groups for Germany, DALYs per 100 g of food *i*, and DALYs for Germany based on the observed diet and a theoretical ideal diet (TMREL).

|  | Diet-related DALYs from two data sources |  | Data points for the linear interpolation |  |  | Results and application of the health indicator (GBD) |  |  | Results and application of the health indicator (SCH) |  |  |
| --- | --- | --- | --- | --- | --- | --- | --- | --- | --- | --- | --- |
| Dietary risk factor/food group<sup>a</sup> | GBD [36] | SCH [18] | $x_i^{Obs}$, mean intake in g/d | $x_i^{TMREL}$, TMREL in g/d [Source] | Diff-erence $x_i^{Obs} - x_i^{TMREL}$ in g/d | DALYs per 100 g of food group $i$ | Observed diet | TMREL diet | DALYs per 100 g of food group $i$ | Observed diet | TMREL diet |
| Diet low in vegetables | 267,977 | 250,022 | 91 | 375 [18] | -284 | -94,490 | -86,360 | -354,337 | -88,159 | -80,574 | -330,595 |
| Diet low in fruits | 325,255 | 239,448 | 154 | 300 [5,50,51] | -146 | -222,444 | -342,077 | -667,331 | -163,760 | -251,832 | -491,281 |
| Diet low in legumes | 189,749 | 245,000 | 5 | 122 [18] | -117 | -162,063 | -7,968 | -197,717 | -209,252 | -10,288 | -255,288 |
| Diet low in nuts and seeds | 74,787 | 428,131 | 5 | 25 [5,52] | -20 | -367,445 | -17,074 | -91,861 | -<br>2,103,493 | -97,742 | -525,873 |
| Diet low in whole grains | 415,671 | 586,353 | 12 | 119 [18] | -107 | -390,079 | -48,523 | -464,194 | -550,252 | -68,447 | -654,800 |
| Diet high in refined grains | n.a. | 18,796 | 222 | 0 [18] | 222 | n.a. | n.a. | n.a. | 8,483 | 18,796 | 0 |
| Diet low in dairy | n.a. | 132,375 | 196<sup>b</sup> | 355 [18] | -159 | n.a. | n.a. | n.a. | -83,408 | -163,724 | -296,099 |
| Diet low in milk | 51,096 | n.a. | 196<sup>b</sup> | 355 [18] | -159 | -32,195 | -63,197 | -114,292 | n.a. | n.a. | n.a. |
| Diet high in eggs | n.a. | 9,398 | 12 | 0 [18] | 12 | n.a. | n.a. | n.a. | 80,965 | 9,398 | 0 |
| Diet low in fish | n.a. | 276,991 | 15 | 131 [18] | -116 | n.a. | n.a. | n.a. | -239,106 | -36,239 | -313,229 |
| Diet high in red meat | 328,075 | 81,679 | 42 | 0 [52] | 42 | 787,702 | 328,075 | 0 | 196,109 | 81,679 | 0 |
| Diet high in processed meat | 380,748 | 274,629 | 52 | 0 [18] | 52 | 726,060 | 380,748 | 0 | 523,699 | 274,629 | 0 |
| Diet high in sugar-sweetened beverages | 74,494 | 339,963 | 144 | 0 [18] | 144 | 51,760 | 74,494 | 0 | 236,213 | 339,963 | 0 |

DALYs = disability-adjusted life years, GBD = Global Burden of Disease, SCH = Schwingshackl, TMREL = theoretical minimum risk exposure level. For ease of presentation, all values are rounded to integers. The reported sums of the dietary risk factors' DALYs (sum of column 2 and sum of column 3) overestimated the diet-related burden slightly compared with the literature, since the latter was adjusted for synergetic effects. For the calculation of observed dietary intakes (column 4), we used the German National Nutrition Survey II (NVS II) (adult population, see Chapter 2.1). For the ideal intakes as TMRELs (column 5), we used only one source of data to keep the data presentation clear. For each food group, we chose the TMREL that best reflects the current proportions between the food groups. Columns 7 and 10 show the results for DALYs per 100 g of food group $i$ (the health indicator) for GBD and SCH data, respectively. Columns 8 and 9 (GBD) and columns 11 and 12 (SCH) show proof-of-concept tests, where we multiplied the health indicator by either $x_i^{Obs}$ or $x_i^{TMREL}$ . <sup>a</sup> Mapping between FoodEx2 food groups and food groups for DALYs is shown in S3 Table. <sup>b</sup> This value includes dairy products and milk.

In **Table 1**, columns 4 and 5 show observed intakes 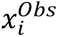 (NVSII) and ideal intakes 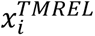 for each food group. The positive and negative values for the differences between observed and ideal intakes in column 6 mathematically represent the food groups for which intakes need to be either increased (e.g., vegetables, which had an observed intake 284 g/d lower than the ideal intake) or decreased (e.g., processed meat, which had an observed intake 52 g/d higher than the ideal intake).

One objective of our study was to unravel whether the calculation of DALYs using linear interpolation was reliable when determining the health impact of various optimized diets. In theory, the sum of DALYs for the ideal diet (TMREL intakes) should 100% offset the sum of DALYs linked to the observed diet, since the implementation of the ideal diet would theoretically eliminate disease burden represented by DALYs. In our case, this would mean a negative number in the same magnitude as the total of 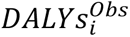 (GBD [36]: 2,107,851 [sum of column 2]; SCH [18]: 2,882,784 [sum of column 3]). Our analysis revealed that similar reductions in DALYs were achieved as reported in both GBD and SCH studies: for GBD, the ideal diet resulted in a 90% decrease of the diet-related disease burden (−1,889,733 DALYs, sum of column 9) and for SCH, the ideal diet resulted in a 99% decrease (−2,876,166 DALYs, sum of column 12).

The sum of calculated DALYs for the observed diet should ideally be 0, which would show that the calculated health impact did not deviate from the sum of DALYs for dietary risks in Germany. For GBD, the observed diet summed up to 218,118 DALYs (sum of column 8), which corresponds to 10% of the total burden reported in the literature [36]. For SCH, the observed diet resulted in 15,618 DALYs (sum of column 11), which corresponds to 1% of the total burden [18].

As an illustration, Fig 1 shows how the observed intake of whole grains (12 g/d, Table 1) is related to the number of DALYs (i.e. 586,353 DALYs for SCH, Table 1); the ideal intake of 119 g/d (Table 1) is related to 0 DALYs. These two data points were used to extrapolate the DALYs related to any given intake (for the formula, see Chapter 2.2). For integration into the optimization model’s database, DALYs per 100 g of a food group (for whole grains, −550,252 DALYs, Table 1) were calculated.

### 3.2 The effects of weighting schemes for health, the environment, and the observed diet on indicator results

Figs 2 a-c shows the results of models with fixed weightings of 35% for health (Fig 2a, models (M) 1-14), the environment (Fig 2b, M15-28), and the observed diet (Fig 2c, M29-42). In each of the modelling series, the weighting of the other components was increased or decreased in 5% increments either from 0% up to 65% or from 65% down to 0%. For each combination, the total sum of weightings was 100%.

**Fig 2.**
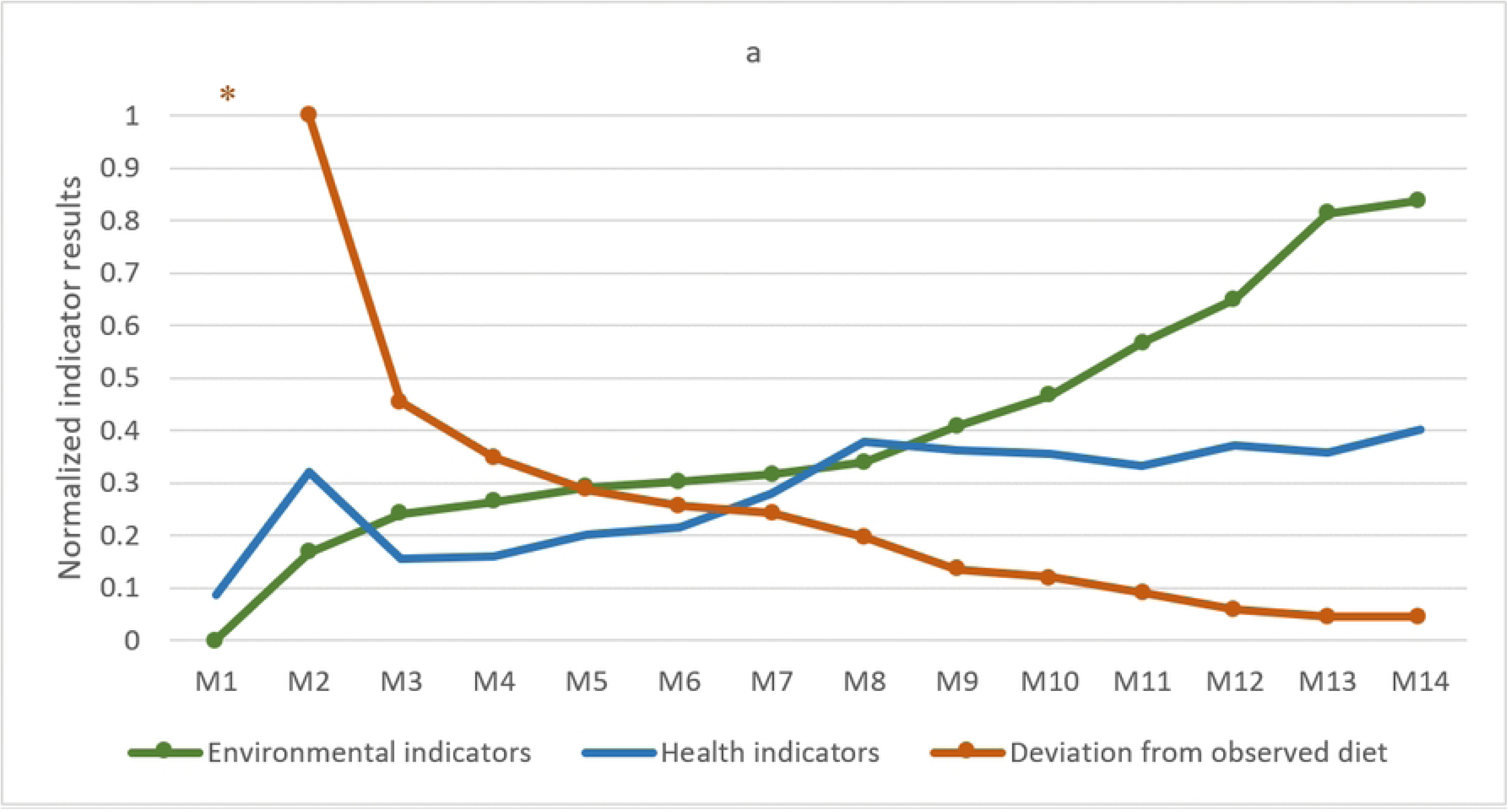
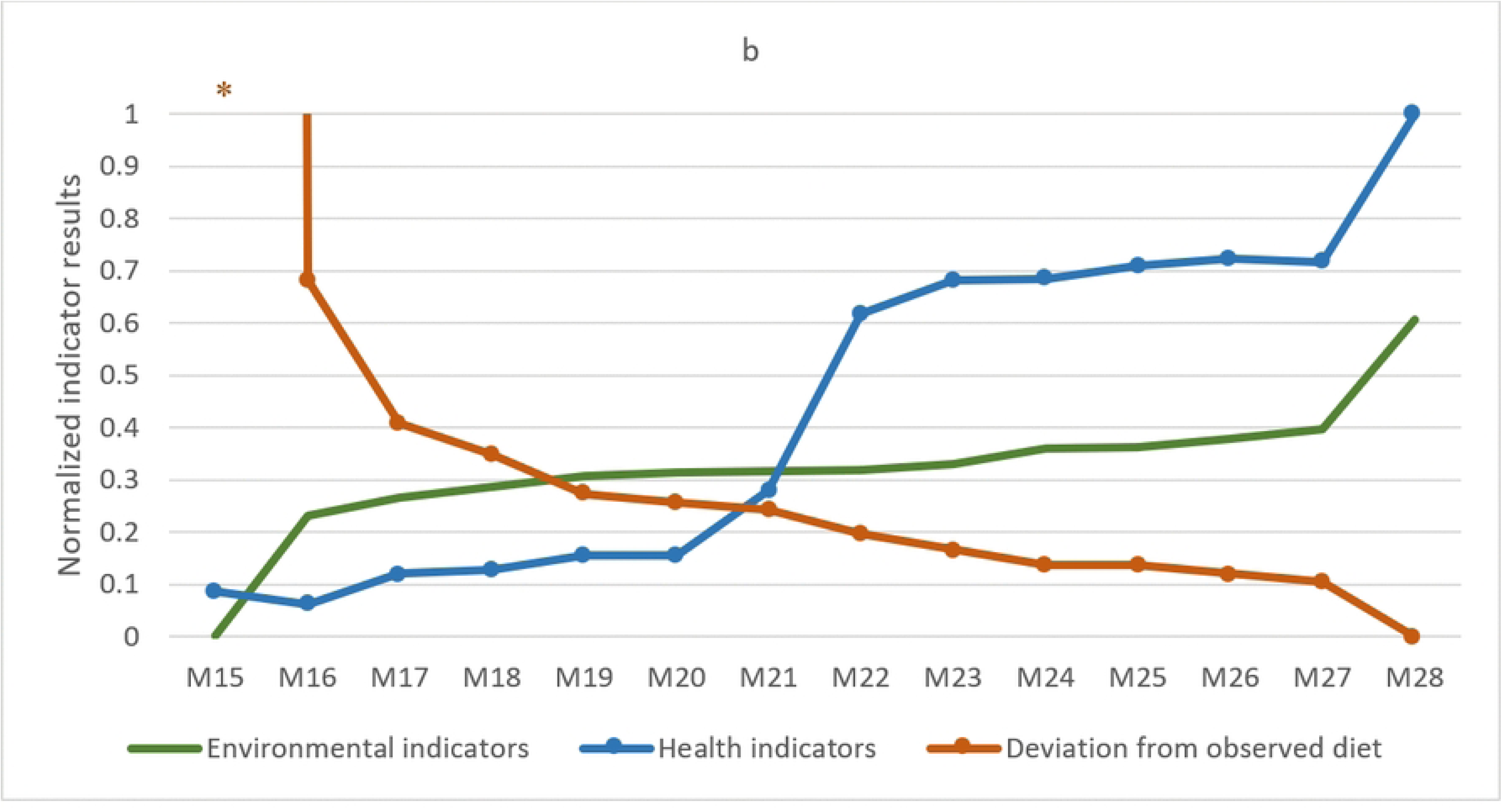
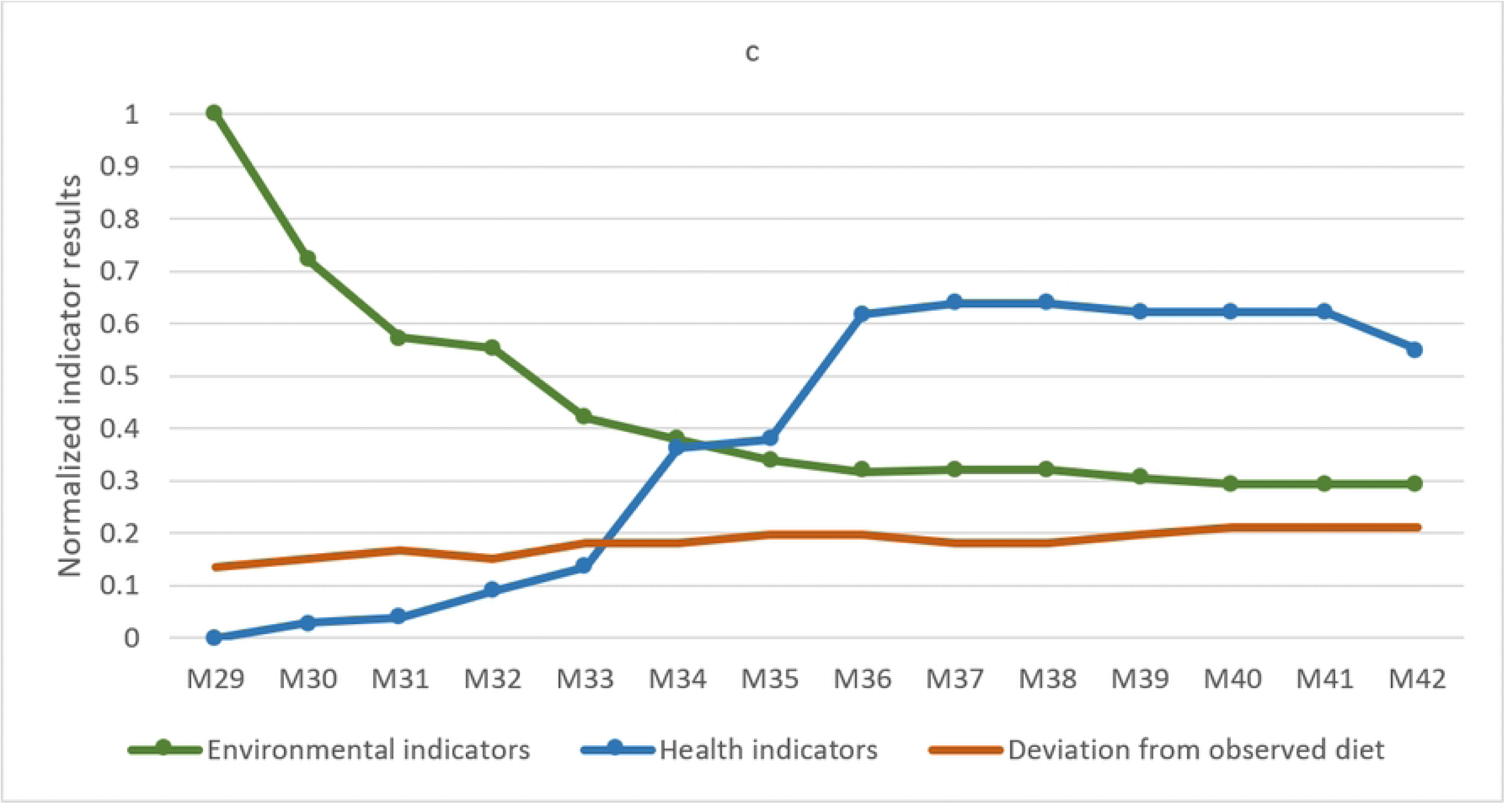
Impact of different weighting schemes of three-objective optimization models on indicator results. For each model (M), one component of the objective function was maintained at a fixed weighting of 35%, while the weighting of the two other components was changed (one increased and one decreased) in increments of 5%: a) Impact of varying weights (5%-steps) on environment (from 65%, M1 to 0%, M14) and deviation from the observed diet (from 0%, M1 to 65%, M14) and a fixed weight on health (35%). b) Impact of varying weights (5%-steps) on health (from 65%, M15 to 0%, M28) and deviation from the observed diet (from 0%, M15 to 65%, M28) and a fixed weight on environment (35%). c) Impact of varying weights (5%-steps) on health (from 65%, M29 to 0%, M42) and environment (from 0%, M29 to 65%, M42) and a fixed weight on deviation from the observed diet (35%). The results are depicted as normalized data (min-max scaling) for ease of presentation. The values from the “observed diet” category for M1 and M15 were not included as part of the min-max scaling. * Values > 35. M = Model. Lines without markings = fixed weighting; lines with dots = varied weightings using 5% increments. The outcomes of different weighting schemes for the three components are depicted as the mean of the reduction in DALYs (GBD and SCH), the mean of the reduction in environmental impact (GHGEs and land use), and deviation from observed dietary intakes for optimized diets. As the aim of the objective function was to minimize each component, smaller values show that the respective objective could be better fulfilled.

Fig 2a (M1-14), in which the health component has a fixed weighting of 35%, shows that although the results for DALYs deviate less than in models where the health component is not fixed (M15–42), there are still variations in the results. A similar trend was seen in Figs 2b and 2c, where weightings for the environment and the observed diet are fixed, respectively, but there is still some deviation as the weightings of other components are changed. This pattern demonstrates the relations between the components, particularly between health and the environment (Figs 2a and 2b). The largest reductions for the environmental impact were achieved in the model with a fixed weight on health and the highest weight (65%) on the environment (M1, Fig 2a), and in the model with a fixed weighting for the environment and the highest weighting (65%) for health (M15, Fig 2b). The largest reduction of DALYs was found in the model with the highest weighting for health (65%) and a fixed weighting for the observed diet (M29, Fig 2c). By contrast, a higher weighting for the observed diet limited the influence of the environment and health components (Figs 2a and 2b) and, therefore, strongly determined the optimization outcomes. The correlations between the indicators for health, the environment, and the observed diet are shown in Table 2.

**Table 2:** Correlations between indicators for health, the environment, and the observed diet.

| Variable 1 | Variable 2 | Pearson's product-moment correlation (95% confidence intervals) of variables 1 and 2 |
| --- | --- | --- |
| Mean of GBD and SCH | Mean of GHGEs (kg CO<sub>2</sub>equivalents/d) and land use (m<sup>2</sup>/year/d) | 0.28 (0.17; 0.39) |
| Mean of GBD and SCH | Mean dietary intakes in g/d | 0.05 (-0.08; 0.17) |
| Mean of GHGEs (kg CO<sub>2</sub>equivalents/d) and land use (m<sup>2</sup>/year/d) | Mean dietary intakes in g/d | -0.07 (-0.19; 0.06) |
| SCH | GBD | 0.72 (0.65; 0.77) |
| Land use (m<sup>2</sup>/year/d) | GHGEs (kg CO<sub>2</sub>equivalents/d) | 0.69 (0.61; 0.75) |

GBD = Global Burden of Disease, SCH = Schwingshackl, GHGEs = greenhouse gas emissions.

We also noted that a steady increase or decrease in weighting was not always associated with a corresponding increase or decrease in indicator results. Fig 2c shows that as the weighting for the environment increased, the reduction of environmental impact stagnated from M36 (35% weighting for the environment) to M42 (65% weighting for the environment). Furthermore, the reduction of DALYs was not necessarily highest with the highest weighting for health. For example, M16 (fixed weighting for the environment, Fig 2b) had a 60% weighting for health, but resulted in a higher reduction of DALYs than M15, where the weighting for health was 5% higher, but the observed diet was not included (0% weighting).

In general, the two-dimensional models (where the weighting was 35% for the fixed dimension and 65% for one of the two varying dimensions) and models with a low weighting (5% or 10%) for one dimension showed the largest differences compared with other models. For example, Figs 2a and 2b clearly indicate that not considering the observed diet generated unstable models, as depicted by the asterisks due to very high differences between the observed diet and the optimized diet (M1 and M15). Furthermore, M1 and M15 had the highest number of food groups at the aggregated FBDG level with optimized quantities of < 1 g/d (12 out of 18 FBDG food groups, plus six discretionary food groups). In the tri-objective models with more balanced weightings, results were more stable (the exceptions were M39 and M40, deviations from the trend in the health indicators, Fig 2c).

In response to changes in component weightings, other indicators, such as the number of food groups with intakes < 1 g/d, and sub-indicators (environment: GHGEs and land use, health: DALYs from SCH and GBD; S4 Table) showed similar trends to those of the results in Figs 2a-c.

### 3.3 Sensitivity analyses for sub-indicators

For the health and environment components, two indicators were integrated into the objective function. Table 3 shows the results of models with different weightings for these indicators, applied within either one-dimensional (100% weighting for health or the environment) or three-dimensional (35% weighting for health and the environment, 30% for the observed diet) objective functions.

**Table 3:**
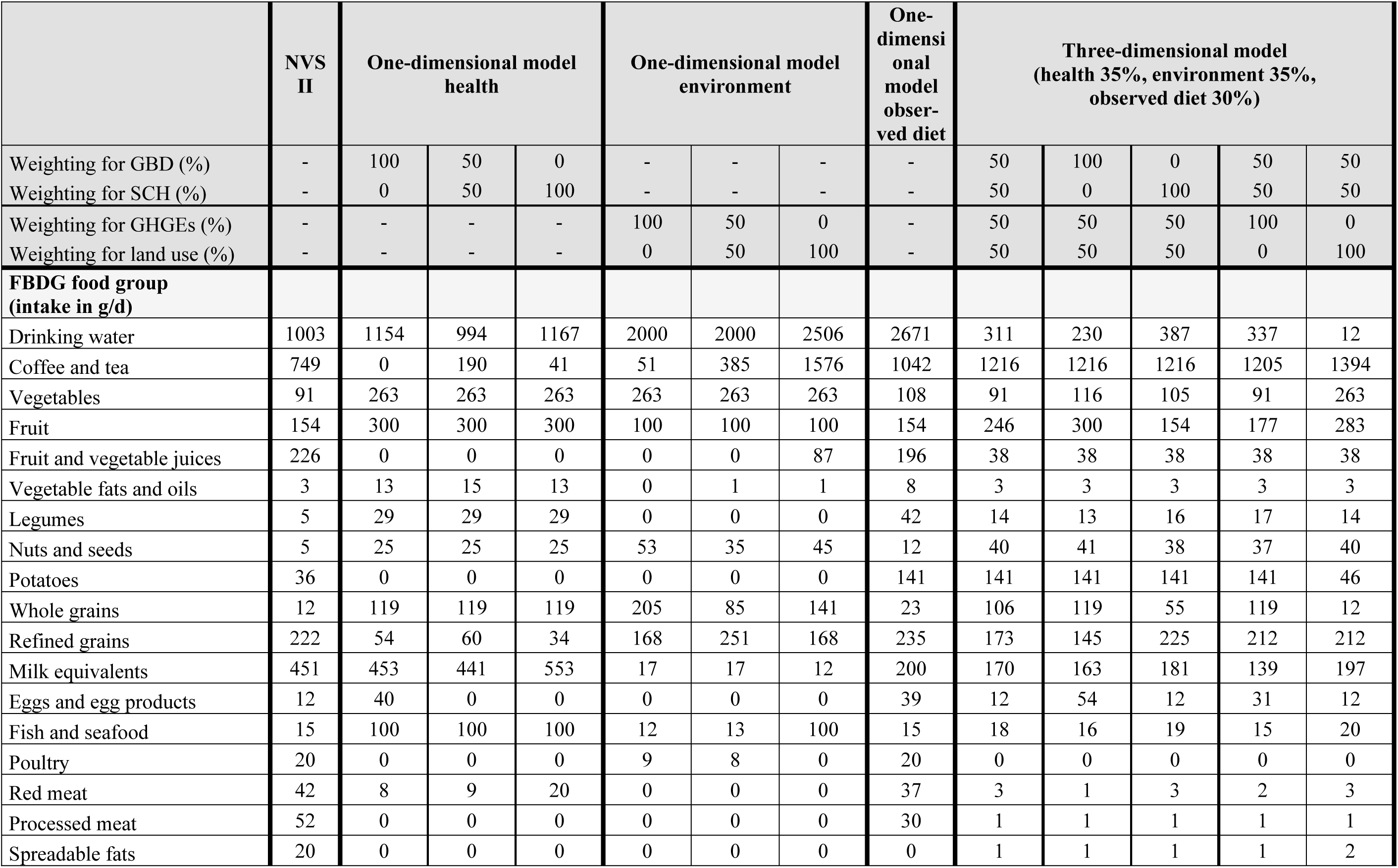
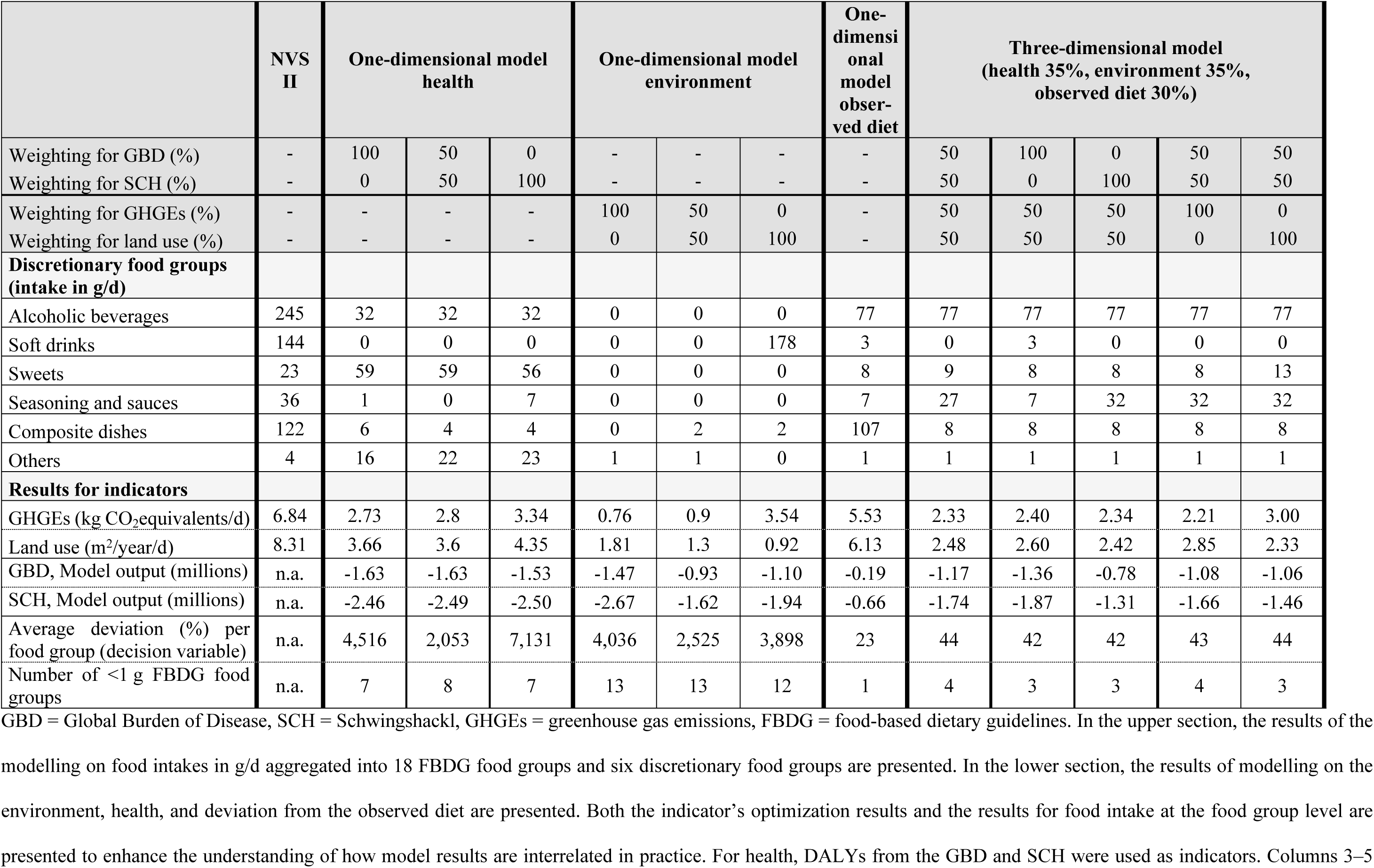
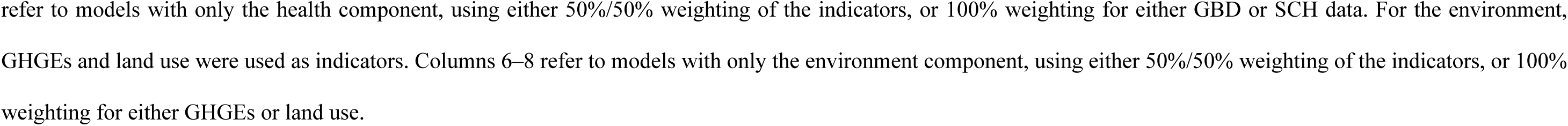
Sensitivity analyses of optimization results investigating different weightings of indicators for health and the environment.

|  | NVS II | One-dimensional model health |  |  | One-dimensional model environment |  |  | One-dimensional model observed diet | Three-dimensional model (health 35%, environment 35%, observed diet 30%) |  |  |  |  |
| --- | --- | --- | --- | --- | --- | --- | --- | --- | --- | --- | --- | --- | --- |
| Weighting for GBD (%) | - | 100 | 50 | 0 | - | - | - | - | 50 | 100 | 0 | 50 | 50 |
| Weighting for SCH (%) | - | 0 | 50 | 100 | - | - | - | - | 50 | 0 | 100 | 50 | 50 |
| Weighting for GHGEs (%) | - | - | - | - | 100 | 50 | 0 | - | 50 | 50 | 50 | 100 | 0 |
| Weighting for land use (%) | - | - | - | - | 0 | 50 | 100 | - | 50 | 50 | 50 | 0 | 100 |
| **FBDG food group (intake in g/d)** |  |  |  |  |  |  |  |  |  |  |  |  |  |
| Drinking water | 1003 | 1154 | 994 | 1167 | 2000 | 2000 | 2506 | 2671 | 311 | 230 | 387 | 337 | 12 |
| Coffee and tea | 749 | 0 | 190 | 41 | 51 | 385 | 1576 | 1042 | 1216 | 1216 | 1216 | 1205 | 1394 |
| Vegetables | 91 | 263 | 263 | 263 | 263 | 263 | 263 | 108 | 91 | 116 | 105 | 91 | 263 |
| Fruit | 154 | 300 | 300 | 300 | 100 | 100 | 100 | 154 | 246 | 300 | 154 | 177 | 283 |
| Fruit and vegetable juices | 226 | 0 | 0 | 0 | 0 | 0 | 87 | 196 | 38 | 38 | 38 | 38 | 38 |
| Vegetable fats and oils | 3 | 13 | 15 | 13 | 0 | 1 | 1 | 8 | 3 | 3 | 3 | 3 | 3 |
| Legumes | 5 | 29 | 29 | 29 | 0 | 0 | 0 | 42 | 14 | 13 | 16 | 17 | 14 |
| Nuts and seeds | 5 | 25 | 25 | 25 | 53 | 35 | 45 | 12 | 40 | 41 | 38 | 37 | 40 |
| Potatoes | 36 | 0 | 0 | 0 | 0 | 0 | 0 | 141 | 141 | 141 | 141 | 141 | 46 |
| Whole grains | 12 | 119 | 119 | 119 | 205 | 85 | 141 | 23 | 106 | 119 | 55 | 119 | 12 |
| Refined grains | 222 | 54 | 60 | 34 | 168 | 251 | 168 | 235 | 173 | 145 | 225 | 212 | 212 |
| Milk equivalents | 451 | 453 | 441 | 553 | 17 | 17 | 12 | 200 | 170 | 163 | 181 | 139 | 197 |
| Eggs and egg products | 12 | 40 | 0 | 0 | 0 | 0 | 0 | 39 | 12 | 54 | 12 | 31 | 12 |
| Fish and seafood | 15 | 100 | 100 | 100 | 12 | 13 | 100 | 15 | 18 | 16 | 19 | 15 | 20 |
| Poultry | 20 | 0 | 0 | 0 | 9 | 8 | 0 | 20 | 0 | 0 | 0 | 0 | 0 |
| Red meat | 42 | 8 | 9 | 20 | 0 | 0 | 0 | 37 | 3 | 1 | 3 | 2 | 3 |
| Processed meat | 52 | 0 | 0 | 0 | 0 | 0 | 0 | 30 | 1 | 1 | 1 | 1 | 1 |
| Spreadable fats | 20 | 0 | 0 | 0 | 0 | 0 | 0 | 0 | 1 | 1 | 1 | 1 | 2 |
| Weighting for GBD (%) | - | 100 | 50 | 0 | - | - | - | - | 50 | 100 | 0 | 50 | 50 |
| Weighting for SCH (%) | - | 0 | 50 | 100 | - | - | - | - | 50 | 0 | 100 | 50 | 50 |
| Weighting for GHGEs (%) | - | - | - | - | 100 | 50 | 0 | - | 50 | 50 | 50 | 100 | 0 |
| Weighting for land use (%) | - | - | - | - | 0 | 50 | 100 | - | 50 | 50 | 50 | 0 | 100 |
| **Discretionary food groups (intake in g/d)** |  |  |  |  |  |  |  |  |  |  |  |  |  |
| Alcoholic beverages | 245 | 32 | 32 | 32 | 0 | 0 | 0 | 77 | 77 | 77 | 77 | 77 | 77 |
| Soft drinks | 144 | 0 | 0 | 0 | 0 | 0 | 178 | 3 | 0 | 3 | 0 | 0 | 0 |
| Sweets | 23 | 59 | 59 | 56 | 0 | 0 | 0 | 8 | 9 | 8 | 8 | 8 | 13 |
| Seasoning and sauces | 36 | 1 | 0 | 7 | 0 | 0 | 0 | 7 | 27 | 7 | 32 | 32 | 32 |
| Composite dishes | 122 | 6 | 4 | 4 | 0 | 2 | 2 | 107 | 8 | 8 | 8 | 8 | 8 |
| Others | 4 | 16 | 22 | 23 | 1 | 1 | 0 | 1 | 1 | 1 | 1 | 1 | 1 |
| **Results for indicators** |  |  |  |  |  |  |  |  |  |  |  |  |  |
| GHGEs (kg CO<sub>2</sub>equivalents/d) | 6.84 | 2.73 | 2.8 | 3.34 | 0.76 | 0.9 | 3.54 | 5.53 | 2.33 | 2.40 | 2.34 | 2.21 | 3.00 |
| Land use (m<sup>2</sup>/year/d) | 8.31 | 3.66 | 3.6 | 4.35 | 1.81 | 1.3 | 0.92 | 6.13 | 2.48 | 2.60 | 2.42 | 2.85 | 2.33 |
| GBD, Model output (millions) | n.a. | -1.63 | -1.63 | -1.53 | -1.47 | -0.93 | -1.10 | -0.19 | -1.17 | -1.36 | -0.78 | -1.08 | -1.06 |
| SCH, Model output (millions) | n.a. | -2.46 | -2.49 | -2.50 | -2.67 | -1.62 | -1.94 | -0.66 | -1.74 | -1.87 | -1.31 | -1.66 | -1.46 |
| Average deviation (%) per food group (decision variable) | n.a. | 4,516 | 2,053 | 7,131 | 4,036 | 2,525 | 3,898 | 23 | 44 | 42 | 42 | 43 | 44 |
| Number of <1 g FBDG food groups | n.a. | 7 | 8 | 7 | 13 | 13 | 12 | 1 | 4 | 3 | 3 | 4 | 3 |

GBD = Global Burden of Disease, SCH = Schwingshackl, GHGEs = greenhouse gas emissions, FBDG = food-based dietary guidelines. In the upper section, the results of the modelling on food intakes in g/d aggregated into 18 FBDG food groups and six discretionary food groups are presented. In the lower section, the results of modelling on the environment, health, and deviation from the observed diet are presented. Both the indicator's optimization results and the results for food intake at the food group level are presented to enhance the understanding of how model results are interrelated in practice. For health, DALYs from the GBD and SCH were used as indicators. Columns 3–5
refer to models with only the health component, using either 50%/50% weighting of the indicators, or 100% weighting for either GBD or SCH data. For the environment, GHGEs and land use were used as indicators. Columns 6–8 refer to models with only the environment component, using either 50%/50% weighting of the indicators, or 100% weighting for either GHGEs or land use.

In the one-dimensional health models, using different weightings for health indicators did not result in large differences between DALYs based on the GBD and SCH. However, using the health dimension alone in models resulted in a considerable change in food intake quantities from the observed diet and reduced the environmental impact by more than 50%.

In a model using only environmental impact as the objective function (columns 6-8) and 50%/50% weighting for GHGEs and land use, GHGE and land use levels of 0.9 kg CO_2_equivalents/d and 1.3 m^2^/year/d were achieved, respectively. When GHGEs alone were used, the outcomes were 0.76 kg CO_2_equivalents/d (GHGEs) and 1.8 m^2^/year/d (land use) vs. 3.54 kg CO_2_equivalents/d (GHGEs) and 0.92 m^2^/year/d (land use) when only land use was applied. Again, the one-dimensional models showed high deviations from the observed diet regarding food intake (up to 4,036%).

In the three-dimensional models (the last five columns), different weightings for GBD and SCH resulted in more obvious differences in health outcomes, with the highest number of spared GBD and SCH DALYs in the 100% GBD model. By contrast, the differences between GHGEs and land use were considerably smaller than in the one-dimensional models. With only GHGEs as the environmental indicator, GHGE and land use levels of 2.21 kg CO_2_equivalents/d and 2.85 m^2^/year/d, respectively, were achieved. In a model with only land use as the environmental indicator, the respective numbers were 3 kg CO_2_equivalents/d and 2.33 m^2^/year/d.

Regarding deviation from the observed diet, the one-dimensional health and environment models showed substantially higher average deviations per food group at the decision variable level (2,053%-2,525%) than the model with only the observed diet as its objective function (23%, column 9). Furthermore, the latter model only had one group out of 24 FBDG food groups with an optimized intake < 1 g/d, whereas in models that did not take the observed diet into account in the objective function eight and 13 food groups had an optimized intake < 1 g/d, respectively.

Additional analyses with the three-dimensional model and further weighting schemes of the indicators (i.e., GHGEs 75% and land use 25%) revealed that once the model included multiple objectives, the impact of each indicator’s weighting decreased (data not shown).

## 4 Discussion

In the present study, we investigated how diet-health relations could be implemented in an optimization model as part of the objective function. We chose DALYs to provide indicators for this purpose because of their comprehensive associations with a broad spectrum of diseases and food groups, as well as the quality and timeliness of the available data. In addition, we analyzed the impact of two different indicators utilizing an easy-to-use algorithm for estimating DALYs for different TMRELs. We showed that the use of two indicators instead of one improved the stability of the model results, although each indicator covered different diseases and methodological approaches. Our interest was further directed to defining the weighting schemes applied to the three-objective function. We observed that a balanced weighting scheme had a stabilizing impact on the model (only marginal differences in the results obtained from the models were found), even when the weighting was incrementally changed. Larger differences were observed if the weighting approached the extremes, which in our case corresponded to a weighting of < 10% in one of three components. Finally, we analyzed how either one- or multidimensional models behaved if one of the two indicators for a component (health or the environment) was removed. When using multiple components under a balanced weighting in the objective function, stable models with low changes per food group and low numbers of food groups set to zero intake were obtained. Under such conditions, the removal of one indicator or exchange of one indicator for another only marginally affected the model output. However, if only one component was used in the objective function, excluding one of the two indicators for this component substantially impacted the model output.

Using linear interpolation, as shown in Fig 1, DALYs can be derived for a continuum of dietary intake assumptions using published data, including future iterations. The introduction of this newly derived health indicator to our optimization model also proved successful, as models using health only or high health weightings achieved the highest reductions in DALYs. A limitation of the linear interpolation is that relations that are actually highly complex are linearized. It can only be seen as an approximation and is not an exact representation of diet-related DALYs. There may be U-shaped risk associations for some diet-disease relations [53], and especially beyond the TMREL, only minor health benefits occur [37]. To account for this limitation, a mechanism to not reduce DALYs beyond the food group’s TMREL was integrated into the model.

In proof-of-concept analyses, we validated our approach by calculating the diet-related burden of disease for Germany and compared this with published results. For total DALYs associated with the observed diet, we derived low positive numbers with the linear interpolation, meaning that the impact of the diet on health was slightly overestimated in the range of 1%-10%. For the TMREL scenario, our estimates reached 90% (GBD) to 99% (SCH) of the published burden, meaning that we underestimated the impact compared with publications based on complex modeling. Such a phenomenon could be related to the flat slope syndrome [54], which occurs if measurement errors, which are inevitable with this kind of approximation, exist. Furthermore, for each food group, we chose only one TMREL from the two possibilities derived from the two sources (mostly from the study published by Schwingshackl et al. [18]). Thus, it is not surprising that our estimated DALYs for the GBD scenario aligned less well with the published data [36] than those from the SCH scenario. The most precise way to account for DALYs would be to define the population attributable fraction/potential impact fraction of a risk factor as a function and to integrate this into the optimization model. However, the exact calculation of DALYs for each TMREL using the population attributable fraction/potential impact fraction requires access to the underlying data used for the calculations and is exclusively tailored to the stated TMREL [37].

In many diet optimization studies, health is often represented by addressing only DRVs [13,19]. To account for relations between dietary intake and health outcomes, constraints are sometimes used to address limits on dietary intake for defined food groups following national FBDGs, e.g., a maximum intake for processed meat [6,15,45,55]. However, this method narrows the solution space, limiting the optimization model’s flexibility to find an optimum for all requirements. The practicality of the simple linear interpolation may encourage other scientists developing FBDGs using diet optimization to include DALYs from diet-health relations into diet optimization studies.

We also analyzed the weighting of components in the objective function (Fig 2). When we incrementally changed the weighting schemes of two components, each model generated different estimates for all three components, independent of whether they had a fixed or incrementally changing weighting. However, the component with a fixed weighting showed less variation across models than components for which weightings increased or decreased. This observation proves that the complex interrelations between components play an important role during model calculations. For example, a high dietary intake of processed meat is associated with relatively high GHGEs as well as high DALYs. Thus, a decrease in processed meat intake in the optimization model due to a high weighting of the environment component also had a positive effect on health. The tendency of higher reductions of DALYs with higher weights for environment was still observed when the weighting on health was fixed (Fig 2a) and vice versa (Fig 2b). This relation was confirmed in one-dimensional models for both health and environment, where significant reductions in both components were observed, despite the relatively low Pearson correlation coefficient between the averages of health and environmental indicators (*R* = 0.28). A synergistic relationship between health and the environment has been frequently reported [56,57], although disparities can occur in specific foods, e.g., fish or milk [58]. This may explain why the estimates of health and environmental impacts in Fig 2c, where health and the environment are weighted against each other and the observed diet is fixed, appear to act in opposition to each other.

By contrast, observed dietary intakes had a very low correlation with the environment (0.07) and health (0.05), and were antagonistic to the environment and health in the weighting analyses. However, without the observed diet component in the one-dimensional models, many food groups were either excluded (e.g., eggs, potatoes, and poultry) or dietary intakes reached the 95^th^ percentile (i.e. 100 g of fish per day). Thereby, such diets become less practical to implement. Then again, when all parameters are combined in a multi-objective model with an observed diet that is far from optimum, trade-offs between the dimensions are less important [58–60]. That the inclusion of the observed diet ensures diet acceptability as well as robust results was also found in a bi-objective optimization model, which used environment and the current diet as components [46]. This suggests that simultaneous consideration of diverse components has a stabilizing effect on models.

Another factor that stabilizes the model is the number of components and their indicators used for the objective function. Our balanced three-dimensional models produced more predictable and consistent outcomes than those where a weighting scheme at the boundaries of the model space was applied. For example, the higher the environment weighting, the lower the environmental impact, in general. By contrast, models where the weighting of one component was very low (i.e., < 5%) or 0 were much more unstable than more balanced weighted three-dimensional models. This phenomenon was further exemplified in the third part of our analysis, which included models with one component only where one of the two indicators was removed (Table 3). Though the health indicators of GBD and SCH had a strong correlation of 0.72, and GHGEs and land use had a strong correlation of 0.69 (analyses used input data from decision variables on FoodEx2 level 3), the exclusion of one indicator led to less stable results. The combination of few objectives to be minimized and the fact that the linear objective function aims to change as few food groups as possible means that few suitable food groups are changed for the stated objective(s) [25]. We found several examples in the literature in which simple optimization models were ideal to answer specific questions, but produced optimized diets that were impractical. Examples of this come from early studies applying diet optimization [61]; simple models that tried to find the optimum for one specific question, e.g., minimum cost [62], GHGEs [45], or deviation from observed diet [63]; and from our own pre-study with a simple model to investigate methodological issues [25]. When it comes to the derivation of realistic dietary patterns, the solution space can be narrowed by using stricter acceptability constraints [64]. However, more restrictive constraints reduce the solution space, which limits the model’s ability to identify the best trade-off among several objectives, or in some cases, to find a solution at all. Excessive constraints can also bias the solution toward an expected outcome through several a priori decisions.

We also noted that changing the weighting of components did not result in predictable changes between model results, such as a gradual decrease in the optimized intake of red meat with a gradual increase in the weighting for the environment. Rather, each adjustment of the model parameters (i.e., component weightings) leads to a new stand-alone model. One explanation for this is that optimization models find one solution along a Pareto front [24]; this front is not linear, but multi-dimensional, like the three-dimensional objective function.

Our findings suggest that in complex, multidimensional models, particularly when indicators are relatively similar in magnitude, the precise weighting of individual indicators may have limited influence on the results. However, in simpler models or cases where there is strong evidence to prioritize specific indicators, weighting decisions can significantly affect the results. With insights into how the three components of our objective function interact with each other, we can formulate approaches to define weighting schemes that achieve the optimal solution of a given problem. A simple approach, if one has no specific target for one or more of the objective function’s components, would be to assign an equal weighting to each component (“balanced”), as done previously for environmental impact and cost [23]. The decision-maker may also assign different weightings according to a component’s significance [24]. However, evidence of the measurement and prioritization of sustainable diet parameters is scarce [65] and, as we have shown in the analyses of the three-dimensional objective function, the components partly act in opposition to each other. Although suggestions for sophisticated mathematical approaches to define weighting schemes exist, all use inherently subjective iterations for solution improvement [24]. Therefore, compromises are an inevitable part of optimal solutions [20,46].

## 5 Conclusions

Our proof-of-concept study demonstrated that the DALY indicator for human health yielded convincing results within the tested framework. Therefore, a novel, easy-to-use method was established to dynamically include diet–health relations within a diet optimization model. The stability of our linear modelling was increased with more components (health, the environment, and the observed diet) and, accordingly, more indicators in the objective function. Fixing the weighting of the observed diet while allowing flexibility in the other dimensions may offer a promising approach for implementation, although this will depend on the dimensions used and their relative importance in specific policy or research contexts. Future work may focus on regularly updating DALYs for relevant food groups; investigating whether the stabilizing function of a multi-dimensional objective function is also obtained with other mathematical approaches, such as quadratic programming; and identifying numerical values for sustainable diet goals to help define the weighting scheme.

## Data Availability

The following data sources were from third parties that can be contacted via the information below: Software: DGE, parts of own developed program solutions can be made available upon request (contact at https://www.dge.de/) MS-Nutrition has provided the basic programming and should be consulted if interested in the program code (contact at https://ms-nutrition.com/en/) Data: Nutrient database: The data on nutrients used for obtaining the results presented in the manuscript are available for the Bundeslebensmittelschlüssel (BLS) from the Max RubnerInstitut (contact: https://blsdb.de/contact) and for LEBTAB from the University of Bonn (https://www.epi.uni-bonn.de/forschung/donald-studie, contact via). Food intake data: The data on food intake underlying the results presented in the study are available from the European Food Safety Authority (EFSA) which has published national consumption data at https://www.efsa.europa.eu/en/data-report/food-consumption-data and can be accessed via a public access per data and documents request (for details, see https://www.efsa.europa.eu/sites/default/files/2023-08/pad-guidance-for-applicants.pdf) Environment data: The data on greenhouse gas emissions and land use is from the SHARP-Indicators Database published in Data in Brief. Instructions to access the dataset are given there. All relevant data on changes applied to the original databases (e.g. assignment of FoodEx2 codes to DALY food groups) are mentioned within the manuscript and its supporting information files.

## Acknowledgments

We thank Arno Lellmann for the thorough compilation of DRV values into a database, Theresa Maria Ting for her contribution to the calculation of the metrics for the deviation of dietary habits, and Ute Alexy for her support with the LEBTAB data. We thank Katja Sandfuchs for her consent to publish the BLS-FoodEx2 matching as personal information. We thank Jan Raphael Schäfer for his support in compiling the database via SQL. We thank Ronja Merschmann for her literature search on multi-objective diet optimization.

## Supporting information

**S1 Table. GBD DALYs.** This table shows the values for DALYs used in this study from the Global Burden of Disease Study.

**S2 Table. Schwingshackl DALYs.** This table shows the values for DALYs used in this study from the study by Schwingshackl et al.

**S3 Table. FoodEx2 DALY allocation.** This table shows the allocation of FoodEx2 codes to FBDG food groups and to according DALYs’ food groups on FoodEx2 level 3.

**S4 Table. Model results and analyses.** This table shows for models 1-42 the optimized dietary intakes and results for environment, health, and deviation from the observed diet.

## References

1. FAO (Food and Agriculture Organization of the United Nations). Food-based dietary guidelines. 2023 [cited 18 Jul 2023]. Available from: https://www.fao.org/nutrition/education/food-based-dietary-guidelines.

2. EFSA (European Food Safety Authority). Scientific opinion on establishing food-based dietary guidelines. EFSA J. 2010; 8:1460. doi: 10.2903/j.efsa.2010.1460.

3. WHO (World Health Organization), editor. Preparation and use of food-based dietary guidelines. Joint FAO/WHO Consultation. Joint FAO/WHO Consultation. ; 1998.

4. Bechthold A, Boeing H, Schwedhelm C, Hoffmann G, Knüppel S, Iqbal K, et al. Food groups and risk of coronary heart disease, stroke and heart failure: a systematic review and dose-response meta-analysis of prospective studies. Crit Rev Food Sci Nutr. 2019; 59:1071–90. doi: 10.1080/10408398.2017.1392288 PMID: 29039970.

5. GBD 2017 Diet Collaborators. Health effects of dietary risks in 195 countries, 1990-2017: a systematic analysis for the Global Burden of Disease Study 2017. Lancet. 2019; 393:1958–72. doi: 10.1016/S0140-6736(19)30041-8 PMID: 30954305.

6. Bechthold A, Boeing H, Tetens I, Schwingshackl L, Nöthlings U. Perspective: food-based dietary guidelines in Europe-scientific concepts, current status, and perspectives. Adv Nutr. 2018; 9:544–60. doi: 10.1093/advances/nmy033 PMID: 30107475.

7. Blomhoff R, Andersen R, Arnesen EK, Christensen JJ, Eneroth H, Erkkola M, et al. Nordic nutrition recommendations 2023. Integrating environmental aspects. Copenhagen: Nordic Council of Ministers; 2023.

8. Crippa M, Solazzo E, Guizzardi D, Monforti-Ferrario F, Tubiello FN, Leip A. Food systems are responsible for a third of global anthropogenic GHG emissions. Nat Food. 2021; 2:198–209. doi: 10.1038/s43016-021-00225-9.

9. Gonzalez Fischer C, Garnett T. Plates, pyramids, and planets. Developments in national healthy and sustainable dietary guidelines: a state of play assessment. Rome: Food and Agriculture Organization of the United Nations; Food Climate Research Network, University of Oxford; 2016.

10. Tetens I, Birt CA, Boeing H, Bodenbach S, Bugel S, de Henauw S, et al. Food-Based Dietary Guidelines - development of a conceptual framework for future food based dietary guidelines in Europe. Report of a FENS Task-Force workshop in Copenhagen, 12-13 March 2018. Br J Nutr. 2020; 124:1338–44. doi: 10.1017/S0007114520002469 PMID: 32624024.

11. FAO (Food and Agriculture Organization), WHO (World Health Organization), editors. Sustainable healthy diets - Guiding principles. Rome; 2019.

12. Gedrich K, Hensel A, Binder I, Karg G. How optimal are computer-calculated optimal diets. Eur J Clin Nutr. 1999; 53:309–18. doi: 10.1038/sj.ejcn.1600727 PMID: 10334657.

13. Gazan R, Brouzes CMC, Vieux F, Maillot M, Lluch A, Darmon N. Mathematical optimization to explore tomorrow’s sustainable diets: a narrative review. Adv Nutr. 2018; 9:602–16. doi: 10.1093/advances/nmy049 PMID: 30239584.

14. Schäfer AC, Schmidt A, Bechthold A, Boeing H, Watzl B, Darmon N, et al. Integration of various dimensions in food-based dietary guidelines via mathematical approaches Report of a DGE/FENS Workshop in Bonn, Germany, 23-24 September 2019. Br J Nutr. 2020:1–18. doi: 10.1017/S0007114520004857 PMID: 33272337.

15. Mertens E, van’t Veer P, Hiddink GJ, Steijns JM, Kuijsten A. Operationalising the health aspects of sustainable diets: a review. Public Health Nutr. 2017; 20:739–57. doi: 10.1017/S1368980016002664 PMID: 27819199.

16. Brink E, van Rossum C, Postma-Smeets A, Stafleu A, Wolvers D, van Dooren C, et al. Development of healthy and sustainable food-based dietary guidelines for the Netherlands. Public Health Nutr. 2019; 22:2419–35. doi: 10.1017/S1368980019001435 PMID: 31262374.

17. GBD 2021 Risk Factors Collaborators. Global burden and strength of evidence for 88 risk factors in 204 countries and 811 subnational locations, 1990-2021: a systematic analysis for the Global Burden of Disease Study 2021. Lancet. 2024; 403:2162–203. doi: 10.1016/S0140-6736(24)00933-4 PMID: 38762324.

18. Schwingshackl L, Knüppel S, Michels N, Schwedhelm C, Hoffmann G, Iqbal K, et al. Intake of 12 food groups and disability-adjusted life years from coronary heart disease, stroke, type 2 diabetes, and colorectal cancer in 16 European countries. Eur J Epidemiol. 2019; 34:765–75. doi: 10.1007/s10654-019-00523-4 PMID: 31030306.

19. van Dooren C. A review of the use of linear programming to optimize diets, nutritiously, economically and environmentally. Front Nutr. 2018; 5:48. doi: 10.3389/fnut.2018.00048 PMID: 29977894.

20. Abejón R, Batlle-Bayer L, Laso J, Bala A, Vazquez-Rowe I, Larrea-Gallegos G, et al. Multi-objective optimization of nutritional, environmental and economic aspects of diets applied to the Spanish context. Foods. 2020; 9:1677. Epub 2020/11/16. doi: 10.3390/foods9111677 PMID: 33207725.

21. Hernández M, Gómez T, Delgado-Antequera L, Caballero R. Using multiobjective optimization models to establish healthy diets in Spain following Mediterranean standards. Oper Res Int J. 2019; 14:2274. doi: 10.1007/s12351-019-00499-9.

22. Soares TF, Escarpinati MC, Gabriel PHR. Multi-objective optimization applied to diet planning for people with diabetes. Pesqui Oper. 2024; 44:e285156. doi: 10.1590/0101-7438.2023.043.00285156.

23. Donati M, Menozzi D, Zighetti C, Rosi A, Zinetti A, Scazzina F. Towards a sustainable diet combining economic, environmental and nutritional objectives. Appetite. 2016; 106:48–57. doi: 10.1016/j.appet.2016.02.151 PMID: 26921487.

24. Madoumier M, Trystram G, Sébastian P, Collignan A. Towards a holistic approach for multi-objective optimization of food processes: a critical review. Trends Food Sci Technol. 2019; 86:1–15. doi: 10.1016/j.tifs.2019.02.002.

25. Schäfer AC, Boeing H, Gazan R, Conrad J, Gedrich K, Breidenassel C, et al. A methodological framework for deriving the German food-based dietary guidelines 2024: Food groups, nutrient goals, and objective functions. PLoS One. 2025; 20:e0313347. Epub 2025/03/12. doi: 10.1371/journal.pone.0313347 PMID: 40073305.

26. Niforou K, Livaniou A, Ioannidou S. FoodEx2 maintenance 2023. EFSA Supporting publications. 2024:EN-8813. doi: 10.2903/sp.efsa.2024.EN-8813.

27. EFSA (European Food Safety Authority). Use of the EFSA comprehensive European food consumption database in exposure assessment. EFSA J. 2011; 9:2097. doi: 10.2903/j.efsa.2011.2097.

28. Merten C, Ferrari P, Bakker M, Boss A, Hearty A, Leclercq C, et al. Methodological characteristics of the national dietary surveys carried out in the European Union as included in the European Food Safety Authority (EFSA) Comprehensive European Food Consumption Database. Food Addit Contam Part A Chem Anal Control Expo Risk Assess. 2011; 28:975–95. doi: 10.1080/19440049.2011.576440 PMID: 21732710.

29. Max Rubner-Institut, Bundesforschungsinstitut für Ernährung und Lebensmittel (MRI). Nationale Verzehrsstudie II. Ergebnisbericht, Teil 2. Die bundesweite Befragung zur Ernährung von Jugendlichen und Erwachsenen. Karlsruhe; 2008.

30. Krems C, Walter C, Heuer T, Hoffmann I. Lebensmittelverzehr und Nährstoffzufuhr - Ergebnisse der Nationalen Verzehrsstudie II. In: Deutsche Gesellschaft für Ernährung e. V., editor. 12. Ernährungsbericht 2012. Bonn; 2012. pp. 40–85.

31. Heuer T, Krems C, Moon K, Brombach C, Hoffmann I. Food consumption of adults in Germany: results of the German National Nutrition Survey II based on diet history interviews. Br J Nutr. 2015; 113:1603–14. Epub 2015/04/13. doi: 10.1017/S0007114515000744 PMID: 25866161.

32. Hartmann B, Vasquez-Calcedo A-L, Bell S, Krems C, Brombach C. The German nutrient database basis for analysis of the nutritional status of the German population. J Food Compost Anal. 2008; 21:S115–S118.

33. Sichert-Hellert W, Kersting M, Chahda C, Schäfer R, Kroke A. German food composition database for dietary evaluations in children and adolescents. J Food Compost Anal. 2007; 20:63–70. doi: 10.1016/j.jfca.2006.05.004.

34. Deutsche Gesellschaft für Ernährung e. V. (DGE), Österreichische Gesellschaft für Ernährung, Schweizerische Gesellschaft für Ernährung, editors. Referenzwerte für die Nährstoffzufuhr. 2nd ed. Bonn; 2021.

35. EFSA (European Food Safety Authority). Overview on tolerable upper intake levels as derived by the Scientific Committee on Food (SCF) and the EFSA Panel on Dietetic Products, Nutrition and Allergies (NDA). 2024 [cited 12 Dec 2024]. Available from: https://www.efsa.europa.eu/sites/default/files/assets/UL_Summary_tables.pdf.

36. Global Burden of Disease Collaborative Network. Global Burden of Disease Study 2021 (GBD 2021) Results. Seattle, United States: Institute for Health Metrics and Evaluation (IHME), 2022 2024 [cited 29 Oct 2024]. Available from: https://vizhub.healthdata.org/gbd-results/.

37. Gorasso V, Morgado JN, Charalampous P, Pires SM, Haagsma JA, Santos JV, et al. Burden of disease attributable to risk factors in European countries: a scoping literature review. Arch Public Health. 2023; 81:116. Epub 2023/06/25. doi: 10.1186/s13690-023-01119-x PMID: 37355706.

38. Plass D, Hilderink H, Lehtomäki H, Øverland S, Eikemo TA, Lai T, et al. Estimating risk factor attributable burden - challenges and potential solutions when using the comparative risk assessment methodology. Arch Public Health. 2022; 80:148. Epub 2022/05/27. doi: 10.1186/s13690-022-00900-8 PMID: 35624479.

39. Mertens E, Kaptijn G, Kuijsten A, van Zanten H, Geleijnse JM, van ‘t Veer P. SHARP-Indicators Database towards a public database for environmental sustainability. Data Brief. 2019; 27:104617. Epub 2019/10/07. doi: 10.1016/j.dib.2019.104617 PMID: 31656843.

40. Mertens E, Kuijsten A, van Zanten HH, Kaptijn G, Dofková M, Mistura L, et al. Dietary choices and environmental impact in four European countries. J Clean Prod. 2019; 237:117827. doi: 10.1016/j.jclepro.2019.117827.

41. Hesselink A, Winkvist A, Lindroos AK, Colombo PE, Bärebring L, Hallström E, et al. High reliance on fortified foods when optimizing diets of adolescents in Sweden for adequate vitamin D intake and climate sustainability. J Steroid Biochem Mol Biol. 2025; 251:106759. Epub 2025/04/07. doi: 10.1016/j.jsbmb.2025.106759 PMID: 40204024.

42. van Dooren C, Tyszler M, Kramer G, Aiking H. Combining low price, low climate impact and high nutritional value in one shopping basket through diet optimization by linear programming. Sustainability. 2015; 7:12837–55. doi: 10.3390/su70912837.

43. Eustachio Colombo P, Patterson E, Schäfer Elinder L, Lindroos AK, Sonesson U, Darmon N, et al. Optimizing school food supply: integrating environmental, health, economic, and cultural dimensions of diet sustainability with linear programming. Int J Environ Res Public Health. 2019; 16:3019. Epub 2019/08/21. doi: 10.3390/ijerph16173019 PMID: 31438517.

44. Fu Y, Irz X. Designing culturally acceptable, nutritious and low environmental impact Finnish diets with mycoprotein: a novel optimization approach. Curr Dev Nutr. 2025; 9:107559. doi: 10.1016/j.cdnut.2025.107559.

45. Ferrari M, Benvenuti L, Rossi L, Santis A de, Sette S, Martone D, et al. Could Dietary Goals and Climate Change Mitigation Be Achieved Through Optimized diet? The experience of modeling the National Food Consumption Data in Italy. Front Nutr. 2020; 7:44. doi: 10.3389/fnut.2020.00048.

46. Bashiri B, Kaleda A, Vilu R. Integrating multi-criteria decision-making with multi-objective optimization for sustainable diet design. J Clean Prod. 2025; 500:145233. doi: 10.1016/j.jclepro.2025.145233.

47. R Core Team. R: A language and environment for statistical computing. Vienna, Austria: 2024 [cited 2 Dec 2024]. Available from: https://www.R-project.org/.

48. Theußl S, Schwendinger F, Hornik K. ROI : an extensible R optimization infrastructure. J Stat Soft. 2020; 94. doi: 10.18637/jss.v094.i15.

49. Berkelaar M. lpSolve: Interface to ‘Lp_solve’ v. 5.5 to Solve Linear/Integer Programs. 2023 [cited 13 Dec 2024]. Available from: https://rdrr.io/cran/lpSolve/.

50. Micha R, Peñalvo JL, Cudhea F, Imamura F, Rehm CD, Mozaffarian D. Association between dietary factors and mortality from heart disease, stroke, and type 2 diabetes in the United States. JAMA. 2017; 317:912–24. doi: 10.1001/jama.2017.0947 PMID: 28267855.

51. Gakidou E, Afshin A, Abajobir AA, Abate KH, Abbafati C, Abbas KM. Global, regional, and national comparative risk assessment of 84 behavioural, environmental and occupational, and metabolic risks or clusters of risks, 1990-2016: a systematic analysis for the Global Burden of Disease Study 2016. The Lancet. 2017; 390:1345–422. doi: 10.1016/s0140-6736(17)32366-8 PMID: 28919119.

52. Fadnes LT, Økland J-M, Haaland ØA, Johansson KA. Estimating impact of food choices on life expectancy: a modeling study. PLoS Med. 2022; 19:e1003889. Epub 2022/02/08. doi: 10.1371/journal.pmed.1003889 PMID: 35134067.

53. Schlesinger S, Neuenschwander M, Schwedhelm C, Hoffmann G, Bechthold A, Boeing H, et al. Food groups and risk of overweight, obesity, and weight gain: a systematic review and dose-response meta-analysis of prospective studies. Adv Nutr. 2019; 10:205–18. doi: 10.1093/advances/nmy092 PMID: 30801613.

54. Magkos F, Manios Y, Babaroutsi E, Sidossis LS. Development and validation of a food frequency questionnaire for assessing dietary calcium intake in the general population. Osteoporos Int. 2006; 17:304–12. Epub 2004/09/10. doi: 10.1007/s00198-004-1679-1 PMID: 15368091.

55. Flynn MAT, O’Brien CM, Ross V, Flynn CA, Burke SJ. Revision of food-based dietary guidelines for Ireland, Phase 2: recommendations for healthy eating and affordability. Public Health Nutr. 2012; 15:527–37. doi: 10.1017/S1368980011002084 PMID: 21914254.

56. Springmann M, Godfray HCJ, Rayner M, Scarborough P. Analysis and valuation of the health and climate change cobenefits of dietary change. Proc Natl Acad Sci USA. 2016; 113:4146–51. doi: 10.1073/pnas.1523119113 PMID: 27001851.

57. Clark MA, Springmann M, Hill J, Tilman D. Multiple health and environmental impacts of foods. Proc Natl Acad Sci USA. 2019; 116:23357–62. doi: 10.1073/pnas.1906908116 PMID: 31659030.

58. Cardinaals RPM, Verly E, Jolliet O, van Zanten HHE, Huppertz T. The complementarity of nutrient density and disease burden for Nutritional Life Cycle Assessment. Front Sustain Food Syst. 2024; 8:1301752. doi: 10.3389/fsufs.2024.1304752.

59. Wilson N, Cleghorn CL, Cobiac LJ, Mizdrak A, Nghiem N. Achieving healthy and sustainable diets: a review of the results of recent mathematical optimization studies. Adv Nutr. 2019; 10, Suppl 4:S389–S403. doi: 10.1093/advances/nmz037 PMID: 31728498.

60. Heerschop SN, Kanellopoulos A, Biesbroek S, van ‘t Veer P. Shifting towards optimized healthy and sustainable Dutch diets: impact on protein quality. Eur J Nutr. 2023; 62:2115–28. Epub 2023/03/23. doi: 10.1007/s00394-023-03135-7 PMID: 36949232.

61. Stigler GJ. The cost of subsistence. J Farm Econom. 1945; 27:303–14.

62. Parlesak A, Tetens I, Dejgård Jensen J, Smed S, Gabrijelčič Blenkuš M, Rayner M, et al. Use of linear programming to develop cost-minimized nutritionally adequate health promoting food baskets. PLoS One. 2016; 11:e0163411. doi: 10.1371/journal.pone.0163411 PMID: 27760131.

63. Darmon N, Ferguson EL, Briend A. A cost constraint alone has adverse effects on food selection and nutrient density: an analysis of human diets by linear programming. J Nutr. 2002; 132:3764–71. doi: 10.1093/jn/132.12.3764 PMID: 12468621.

64. Horgan GW, Perrin A, Whybrow S, Macdiarmid JI. Achieving dietary recommendations and reducing greenhouse gas emissions: modelling diets to minimise the change from current intakes. Int J Behav Nutr Phys Act. 2016; 13:46. doi: 10.1186/s12966-016-0370-1 PMID: 27056829.

65. Jones AD, Hoey L, Blesh J, Miller L, Green A, Shapiro LF. A systematic review of the measurement of sustainable diets. Adv Nutr. 2016; 7:641–64. doi: 10.3945/an.115.011015 PMID: 27422501.

